# Predictors of Mortality among Paediatric Snakebite Patients in Northeastern Nigeria: A Survival Analysis Study

**DOI:** 10.1101/2025.08.08.25333277

**Authors:** Aashna Uppal, Nicholas Amani Hamman, Nuhu Mohammed, Abigail Micah Umar, M Surya, Eric Nyarko, Suraj Abdulkarim, Trudie Lang, Joshua Abubakar Difa

## Abstract

**Background:** Paediatric patients of snakebite envenoming are particularly vulnerable to severe complications, however, risk factors for mortality remain largely unexamined in pediatric populations, particularly within rural Nigerian settings, where snakebite envenoming is prevalent.

**Methods:** A retrospective study was conducted using medical records at the Snakebite Treatment and Research Hospital (SBTRH) in Northeastern Nigeria, including paediatric patients aged 0 through 17 who presented with a snakebite from January through December 2024. We used descriptive statistics to summarize patient characteristics grouped by outcome at the end of hospitalization (death versus discharge alive). Subsequently, we estimated cumulative survival probability over time using Kaplan-Meier curves and identified predictors of mortality using multivariable Cox proportional hazards models.

**Results:** SBTRH managed 2,192 snakebite patients between 1 January and 31 December 2024, out of which 723 paediatric patients were included in analyses; 702 of these patients were discharged alive, while 21 died in hospital. Those who took four hours or more to arrive at the hospital after being bitten were more likely to die at hospital discharge compared to those who arrived in less than four hours (hazard ratio (HR) = 5.87, 95% CI = 1.15-29.95). Further, those who were not given antivenom were more likely to die at hospital discharge than those who were given antivenom (HR = 10.07, 95% CI = 3.89-26.07).

**Conclusion:** This study identifies delayed hospital presentation and lack of antivenom administration among key predictors of in-hospital mortality among paediatric snakebite patients in Northeastern Nigeria. These findings underscore the need for urgent public health interventions, including the strengthening of antivenom supply chains, eliminating financial barriers to treatment, and the implementation of targeted educational campaigns aimed at early care-seeking behaviour.

## Introduction

Snakebite envenoming (SBE) persists as a critically under-recognized public health crisis, inflicting profound suffering and loss of life, especially among the rural and underserved populations of Sub-Saharan Africa (SSA) (1,2,3,4). The World Health Organization (WHO) estimates that globally, snakebites affect 5.4 million people annually, of which about 2.7 million people suffer from envenomation, and approximately 81,000 to 138,000 die (5). This translates to approximately 10 snakebites per minute, with around 5 resulting in envenomation, and one death occurring every 4 to 7 minutes. Nigeria has the highest incidence of snakebite in SSA, with most cases occurring in the northern savannah and Benue Valley regions, where farming, herding, and rural lifestyles increase the risk of snake encounters (6,7,8). Despite the scale of the problem in this region, snakebite remains under-reported, poorly studied, and inadequately addressed by health systems, especially when it comes to its impact on vulnerable populations such as children (9,10).

In northeastern Nigeria, the saw-scaled viper (*Echis ocellatus*), the puff adder (*Bitis arietans*), and cobras (*Naja* spp.) are the three predominant venomous snake species reported, collectively responsible for the majority of SBE cases in the region (6,11). Among them, the saw-scaled viper is the most frequently implicated and is associated with severe clinical outcomes such as coagulopathy, systemic hemorrhage, hypovolemic shock, tissue necrosis, and, in many cases, death (12). The puff adder and cobras contribute significantly as well, with envenomation often resulting in cytotoxic and neurotoxic effects (13, 14, 15).

Paediatric patients are particularly vulnerable to severe complications due to their smaller body size, lower physiological reserves, and dependence on adults for timely medical attention, which often results in delayed access to care (16, 17, 18). Risk factors such as delayed hospital presentation, late or insufficient antivenom administration, the species of snake involved, the anatomical site and time of the bite, and the presence of systemic complications (for example, shock, coagulopathy, or central nervous system involvement) have been associated with mortality in studies including adult patients (11,19,20,21). However, these risk factors remain largely unexamined in pediatric populations, particularly within rural Nigerian settings, using time-sensitive analytics. This highlights the urgent need for context-specific research and the use of the most robust analytical methods to understand better the time-sensitive nature of snakebite outcomes in this high-risk group.

This study aims to address these gaps by conducting a one-year retrospective survival analysis of pediatric snakebite patients admitted to the Snakebite Treatment and Research Hospital (SBTRH) in Gombe State, northeastern Nigeria. Specifically, the study will identify clinical and demographic factors associated with an increased mortality risk among pediatric patients, quantify the impact of treatment-related variables, such as antivenom cost, dosage, and timing, on survival outcomes, and evaluate the role of delays in presentation. By integrating methodological rigor with locally relevant clinical data, this study aims to generate actionable insights into the epidemiology and outcomes of pediatric SBE in Northeastern Nigeria. The findings will help inform more effective triage protocols, optimize the use of antivenom, and improve access to life-saving interventions, ultimately contributing to a reduction in mortality and morbidity among one of the most vulnerable patient groups.

## Materials and Methods

The study setting and patient inclusion criteria have been described elsewhere (23). In sum, this was a retrospective study conducted at SBTRH in Kaltungo, Gombe State, Northeastern Nigeria. Paediatric patients aged 0 through 17 who presented at SBTRH with a snakebite from January through December 2024, who did not abscond or leave against medical advice, were included in the study. Patient folders were accessed by trained medical staff at SBTRH (authors NAH, NM, AMU) on 1 May 2025; these staff members could identify individual patients during data collection but ensured that no identifiable data was documented for the purposes of research. Specifically, the following information was documented in a Microsoft Excel file from each included patient folder: month of snakebite occurrence, patient age, patient sex, patient state of origin, patient occupation, anatomical site of snakebite, species of snake, number of antivenom vials used, cost of antivenom (free vs. paid), number of hours between snakebite and presentation to hospital, number of hours between presentation to hospital and antivenom administration, duration of hospital stay and patient outcome (death versus discharged alive).

Patient data was then cleaned and analyzed using R software (version 4.3.1). We performed a three-stage analysis. First, we used descriptive statistics (proportions, medians, and interquartile ranges) to summarize patient characteristics grouped by outcome at the end of hospitalization (death versus discharge alive). The student’s t-test was used for continuous variables, Fisher’s exact test was used for categorical variables with cell counts less than five, and Pearson’s Chi-squared test was used for categorical variables with cell counts greater or equal to five, to summarize differences in patient characteristics between those who died in hospital and those who were discharged alive. Variables for which p-values for the corresponding statistical test were less than 0.05 were considered to be significantly associated with patient mortality. To prepare for the following analyses, we created a subset of patients with complete information and excluded any variable categories with zero cell counts.

Second, we estimated cumulative survival probability over time using Kaplan-Meier curves. We estimated overall survival for the paediatric cohort, as well as group-wise survival for each variable, separately. Variables that had p-values less than 0.05 in these analyses were considered to be significantly associated with better or worse survival. Further, we considered three distinct time-to-event measures: (1) days between snakebite and hospital discharge, (2) days between hospital arrival and discharge, and (3) days between antivenom administration and discharge. Notably, for the third time-to-event measure, only patients who received antivenom were included in the analyses.

Third, we identified predictors of death using multivariable Cox proportional hazards models. We included only variables that did not violate the proportional hazards assumption. Further, if variables demonstrated high degrees of multicollinearity, separate models were created for each of the collinear variables. Through this process, adjusted hazard ratios were produced. Hazard ratios with 95% confidence intervals (CIs) that did not overlap one indicated a significant increase (above one) or decrease (below one) in mortality risk. Similar to the Kaplan-Meier curves, cox proportional hazards modelling was performed for each of the three time-to-event measures.

Ethical clearance was obtained from the Gombe State Ministry of Health (Ref: MOH/ADM/621/V.1/497), and necessary approvals were obtained from the Gombe State Hospital Services Management Board. This study was part of a larger project to generate evidence from existing health data at SBTRH. Through this approval process, the requirement for individual patient consent was waived.

## Results

SBTRH managed 2,192 snakebite patients between 1 January and 31 December 2024, out of which 723 paediatric patients were included in analyses; 702 of these patients were discharged alive, while 21 died in hospital. The characteristics of these patients are described in Table 1.

The median age of patients was 12 years old (interquartile range 8 to 15 years), with the largest proportion of patients belonging to the 10 to 14 year age group (n = 300, 41%). Two-thirds of patients were male (n = 480, 66%) and took four hours or more to arrive at the hospital (n = 468, 65%). Most patients were prescribed a single dose of antivenom (n = 512, 71%) one hour or more after hospital admission (n = 491, 68%). Of note, 112 patients did not receive antivenom due to a lack of availability at the time. Approximately half of the cohort received antivenom free of charge (n = 351, 49%) and arrived at the hospital from within Gombe state (n = 364, 50%). Snakebites demonstrated a seasonal trend, with one-quarter of the whole year’s bites occurring in March and April (n = 183, 25%), at the onset of the rainy season. More than half of the cohort was under care (n = 413, 57%), which refers to patients aged 16 years and under who are neither working nor in school. Most bites occurred in the upper limb (n = 434, 60%), and the majority of patients were bitten by carpet vipers (n = 559, 77%).

There were notable differences in patient characteristics between those who died in the hospital and those who were discharged alive. The number of hours between bite and hospitalization (p = 0.012), antivenom dose (p < 0.001), hours between hospitalization and antivenom administration (p = 0.002), antivenom cost (p < 0.001), and state of origin (p = 0.024) were all significantly associated with survival.

**Table 1.** Patient Characteristics.

| Variable | Overall, N = 723<sup>1</sup> | Patient Outcome at Discharge |  | p-value<sup>2</sup> |
| --- | --- | --- | --- | --- |
|  |  | Dead, N = 21<sup>1</sup> | Alive, N = 702<sup>1</sup> |  |
| Patient Demographics |  |  |  |  |
| Age | 12.0 (8.0, 15.0) | 14.0 (8.0, 16.0) | 11.5 (8.0, 15.0) | 0.209 |
| Age Group |  |  |  | 0.171 |
| 10 to 14 | 300 | 7 (2.3%) | 293 (98%) |  |
| 5 to 9 | 189 | 3 (1.6%) | 186 (98%) |  |
| 15 to 17 | 185 | 8 (4.3%) | 177 (96%) |  |
| 0 to 4 | 49 | 3 (6.1%) | 46 (94%) |  |
| Sex |  |  |  | 0.978 |
| Male | 480 | 14 (2.9%) | 466 (97%) |  |
| Female | 243 | 7 (2.9%) | 236 (97%) |  |
| State of Origin |  |  |  | 0.024 |
| Gombe | 364 | 9 (2.5%) | 355 (98%) |  |
| Taraba | 122 | 1 (0.8%) | 121 (99%) |  |
| Adamawa | 90 | 4 (4.4%) | 86 (96%) |  |
| Bauchi | 67 | 1 (1.5%) | 66 (99%) |  |
| Borno | 50 | 6 (12%) | 44 (88%) |  |
| Yobe | 26 | 0 (0%) | 26 (100%) |  |
| Other<sup>4</sup> | 4 | 0 (0%) | 4 (100%) |  |
| Occupation |  |  |  | 0.499 |
| Under Care<sup>5</sup> | 413 | 10 (2.4%) | 403 (98%) |  |
| Farmer | 143 | 7 (4.9%) | 136 (95%) |  |
| Student | 141 | 4 (2.8%) | 137 (97%) |  |
| House Wife | 21 | 0 (0%) | 21 (100%) |  |
| Other<sup>4</sup> | 5 | 0 (0%) | 5 (100%) |  |
| Circumstances of Snakebite |  |  |  |  |
| Month of Snakebite Occurrence |  |  |  |  |
| April | 102 | 3 (2.9%) | 99 (97%) |  |
| March | 81 | 0 (0%) | 81 (100%) |  |
|  |  | Dead, N = 21<sup>1</sup> | Alive, N = 702<sup>1</sup> |  |
| August | 73 | 4 (5.5%) | 69 (95%) |  |
| October | 72 | 4 (5.6%) | 68 (94%) |  |
| July | 67 | 3 (4.5%) | 64 (96%) |  |
| May | 64 | 0 (0%) | 64 (100%) |  |
| June | 59 | 1 (1.7%) | 58 (98%) |  |
| September | 58 | 3 (5.2%) | 55 (95%) |  |
| February | 47 | 1 (2.1%) | 46 (98%) |  |
| November | 39 | 2 (5.1%) | 37 (95%) |  |
| January | 31 | 0 (0%) | 31 (100%) |  |
| December | 30 | 0 (0%) | 30 (100%) |  |
| **Hours Between Bite and Hospitalization<sup>3</sup>** |  |  |  | 0.012 |
| ≥ 4 hours | 468 | 19 (4.1%) | 449 (96%) |  |
| < 4 hours | 255 | 2 (0.8%) | 253 (99%) |  |
| **Site of Snakebite** |  |  |  | 0.361 |
| Upper Limb | 434 | 10 (2.3%) | 424 (98%) |  |
| Lower Limb | 284 | 11 (3.9%) | 273 (96%) |  |
| Other<sup>4</sup> | 5 | 0 (0%) | 5 (100%) |  |
| **Snake Species** |  |  |  | 0.831 |
| Carpet Viper | 559 | 16 (2.9%) | 543 (97%) |  |
| Unidentified | 157 | 5 (3.2%) | 152 (97%) |  |
| Other<sup>4</sup> | 7 | 0 (0%) | 7 (100%) |  |
| **Treatment** |  |  |  |  |
| **Antivenom Dose** |  |  |  | <0.001 |
| 1 vial | 512 | 11 (2.1%) | 501 (98%) |  |
| 0 vials | 112 | 10 (8.9%) | 102 (91%) |  |
| ≥ 2 vials | 99 | 0 (0%) | 99 (100%) |  |
| **Hours Between Hospitalization and Antivenom Administration<sup>3</sup>** |  |  |  | 0.002 |
| ≥ 1 hour | 491 | 10 (2.0%) | 481 (98%) |  |
| None given | 112 | 10 (8.9%) | 102 (91%) |  |
| < 1 hour | 87 | 1 (1.1%) | 86 (99%) |  |
| Not Reported | 33 | 0 | 33 |  |
| **Antivenom Cost** |  |  |  | <0.001 |
| Free | 351 | 4 (1.1%) | 347 (99%) |  |
| Paid<sup>6</sup> | 260 | 7 (2.7%) | 253 (97%) |  |
| None given | 112 | 10 (8.9%) | 102 (91%) |  |

<sup>1</sup> n (%) or median (interquartile range)
<sup>2</sup> Student's t-test for continuous variables; Fisher's exact test for categorical variables (cell counts less than five); Pearson's Chi-squared test for categorical variables (cell counts greater or equal to five). For one variable (month of presentation), p-values could not be calculated due to low cell counts.
<sup>3</sup> The median number of hours between bite and hospitalization is 4, and the median number of hours between hospitalization and antivenom administration is 1.
<sup>4</sup> Other values for state of origin are Cameroun and Plateau, for occupation are applicant and business, for site of snakebite are back and neck, and for snake species are cobra, night adder, and puff adder.
<sup>5</sup> Under care refers to patients 16 years of age and under who are neither working nor in school.
<sup>6</sup> Paid means the patient paid either for a full or partial dose.

Figure 1 illustrates the overall survival curve for a subset of patients from this cohort with no missing information and variable cell counts above zero (n = 677). The survival probability remained high throughout the time period (days between snakebite and hospital discharge), decreasing from 100% to just above 88%. Results for overall survival from hospital admission to discharge (Figure S1) and from antivenom administration to discharge (Figure S2) were qualitatively similar, albeit with higher survival probabilities.

**Fig 1.**
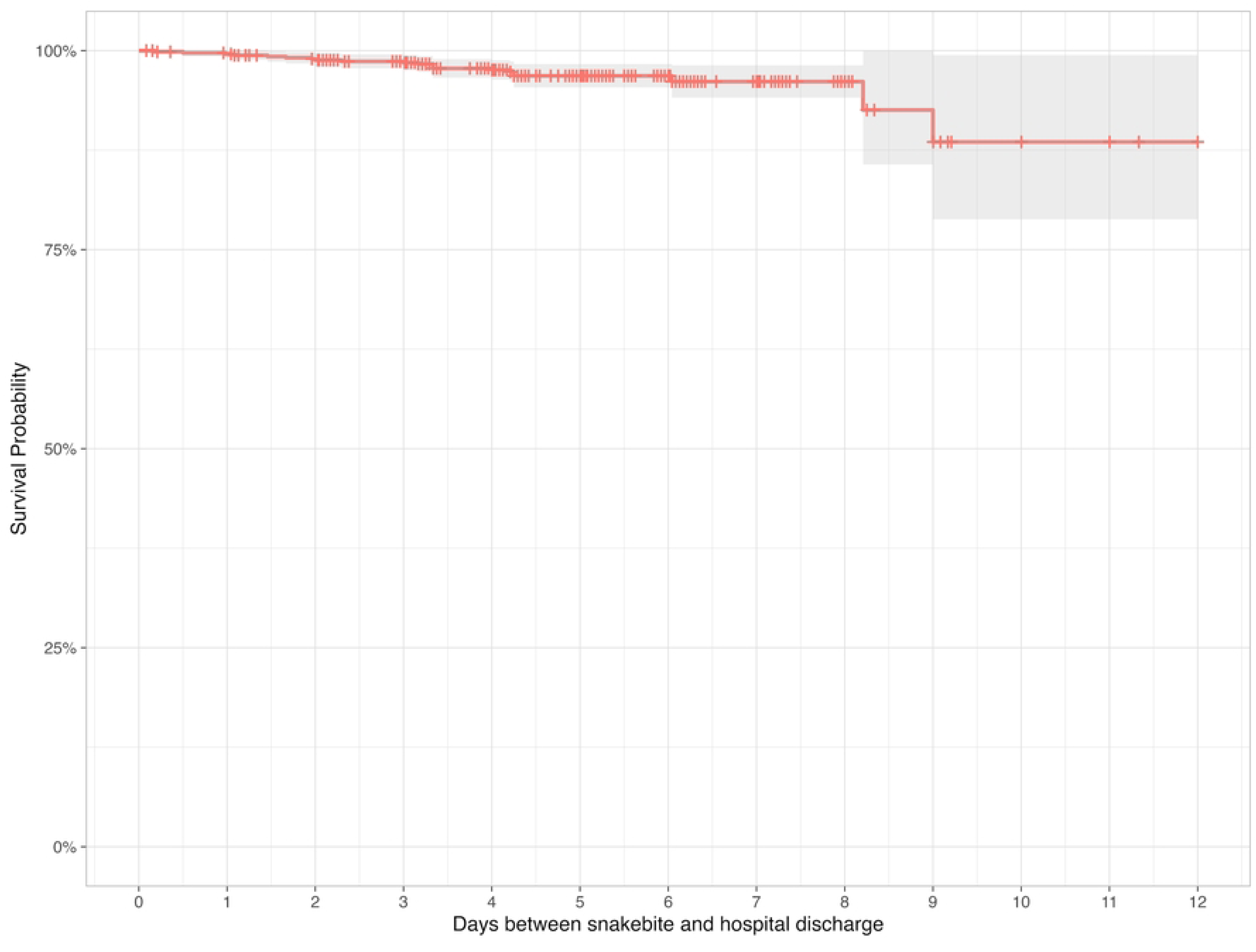
Overall survival probability from snakebite to hospital discharge

Figures 2-6 show Kaplan-Meier curves for covariates that were significantly associated with survival in univariate analyses. Antivenom administration (p < 0.0001), antivenom cost (p < 0.0001), and hours between hospital admission and antivenom administration (p < 0.0001) were all significantly associated with cumulative survival probability, illustrating that those who had antivenom had consistently higher probability of survival than those who did not receive antivenom (Figures 2, 3, and 4). The hours between bite and hospitalization also demonstrated a significant association with cumulative survival probability (p = 0.043); those who arrived four hours or more after being bitten had a worse survival rate than those who arrived in less than four hours (Figure 5). Finally, seasonality demonstrated a significant association with cumulative survival probability (p = 0.045), with snakebites occurring during the rainy season having worse survival than those occurring during the dry season (Figure 6). Other covariates did not demonstrate a significant association with the cumulative survival probability between snakebite and hospital discharge (Figures S3-S8).

**Fig 2.**
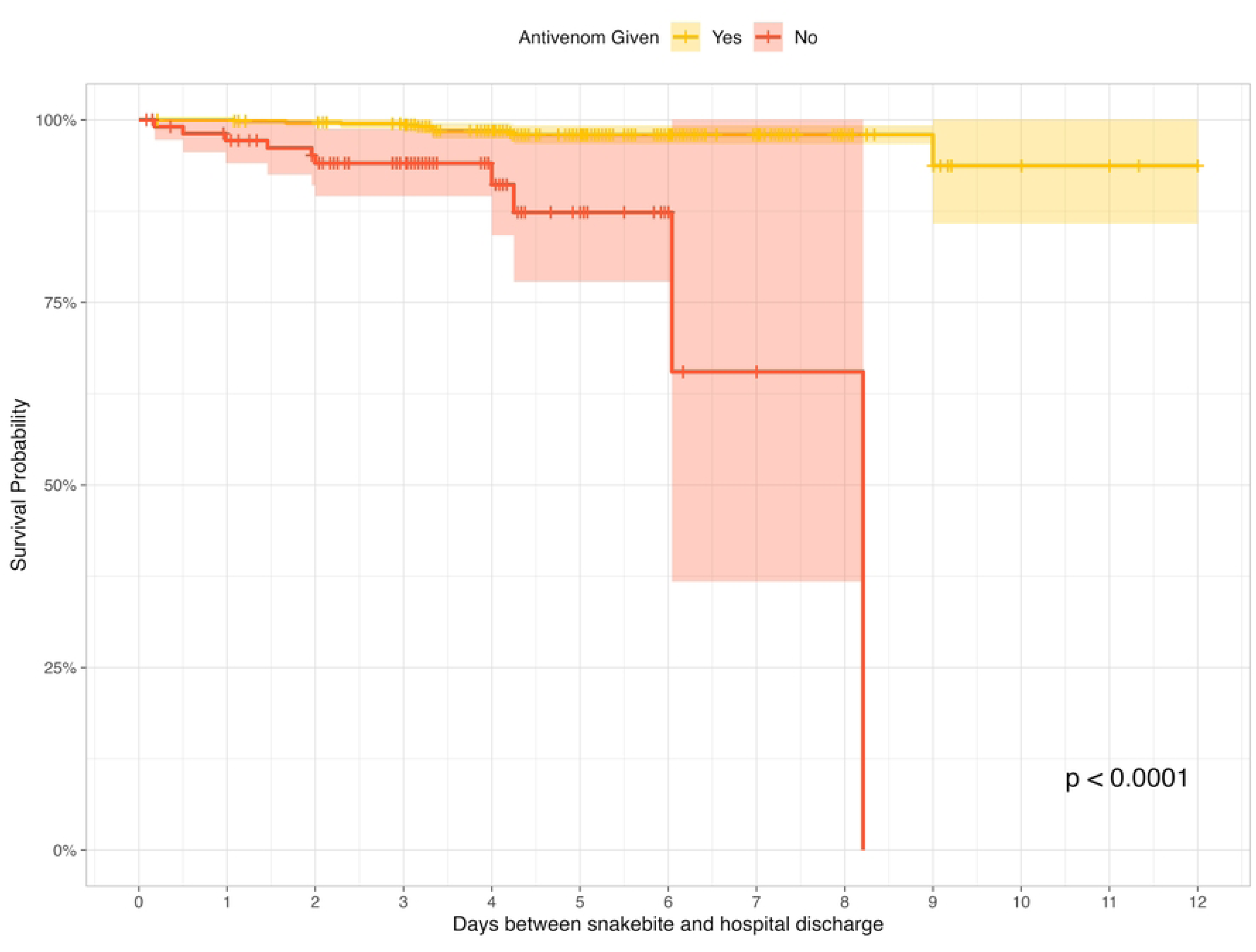
Cumulative survival probability by antivenom administration from bite to hospital discharge

**Fig 3.**
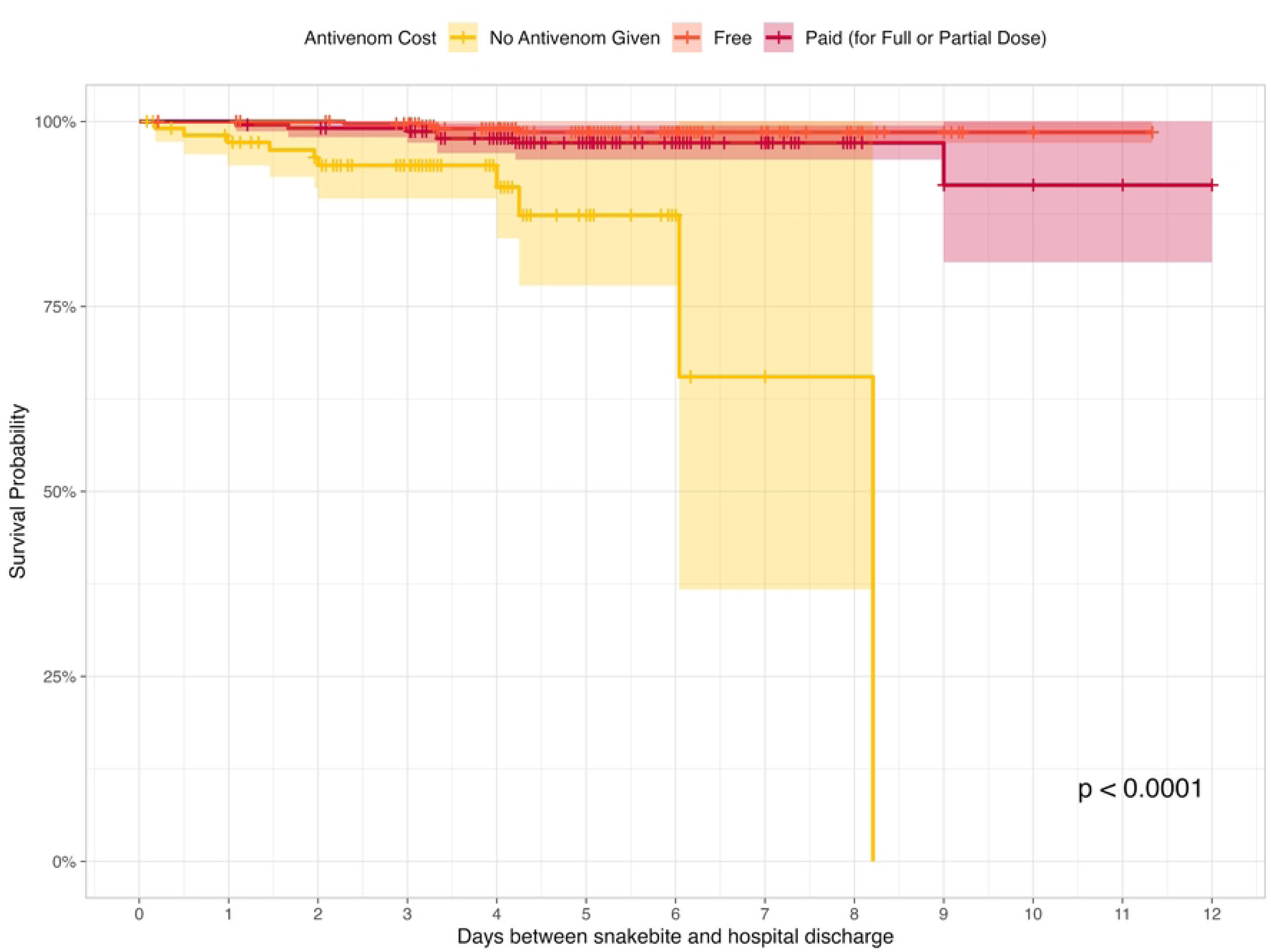
Cumulative survival probability by antivenom cost from bite to hospital discharge

**Fig 4.**
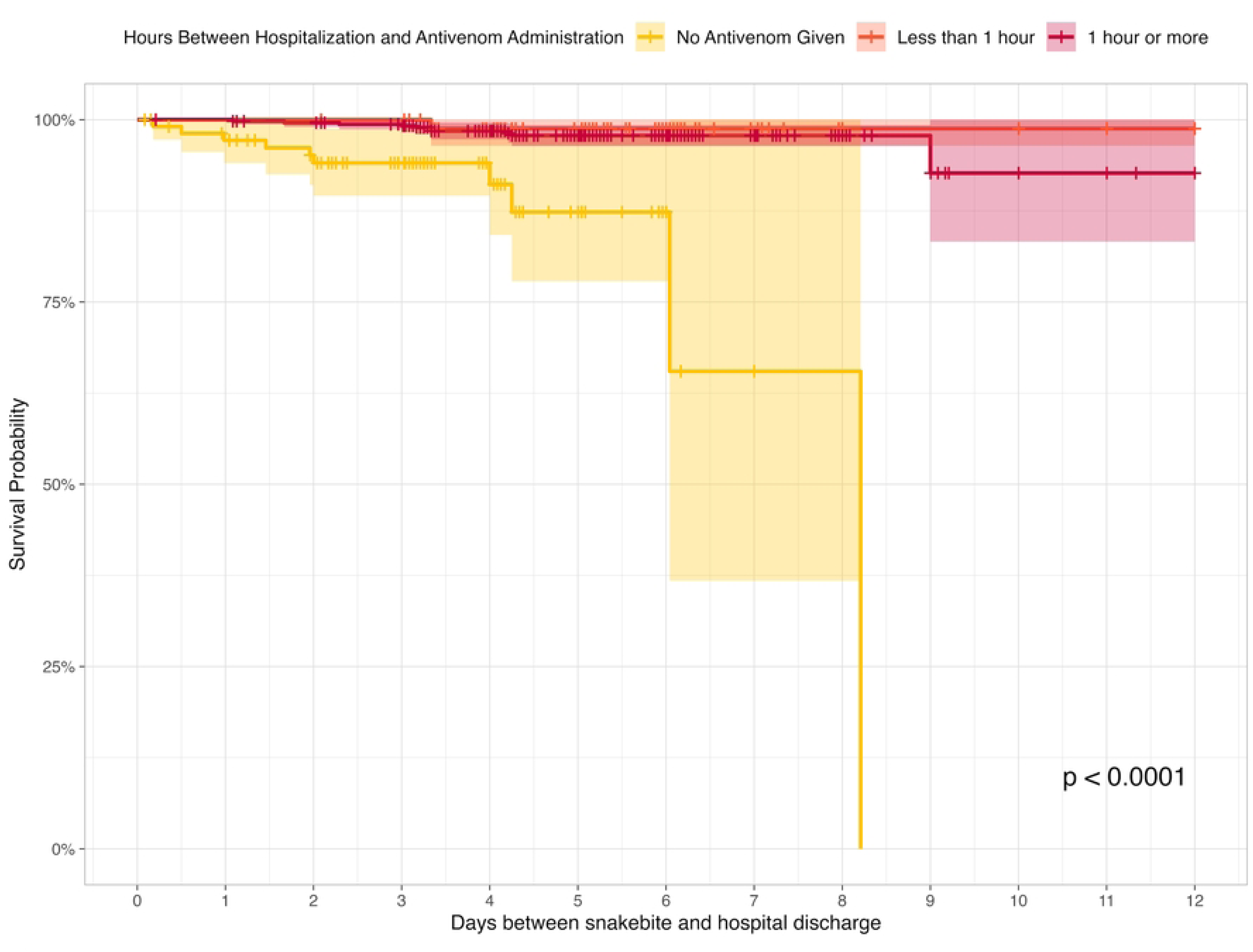
Cumulative survival probability by time hours between hospital admission and antivenom administration from bite to hospital discharge

**Fig 5.**
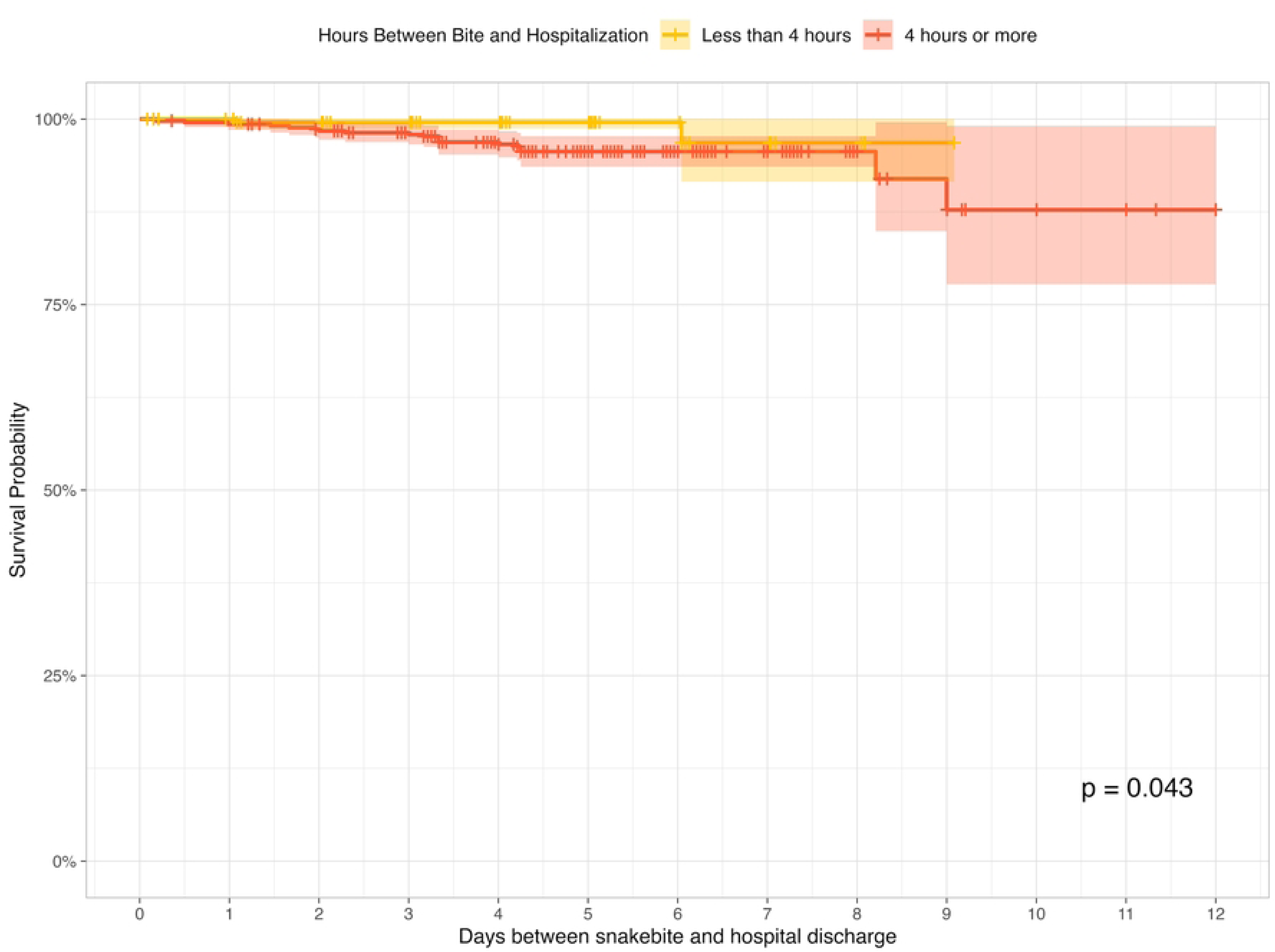
Cumulative survival probability by time between bite and hospitalization from bite to hospital discharge

**Fig 6.**
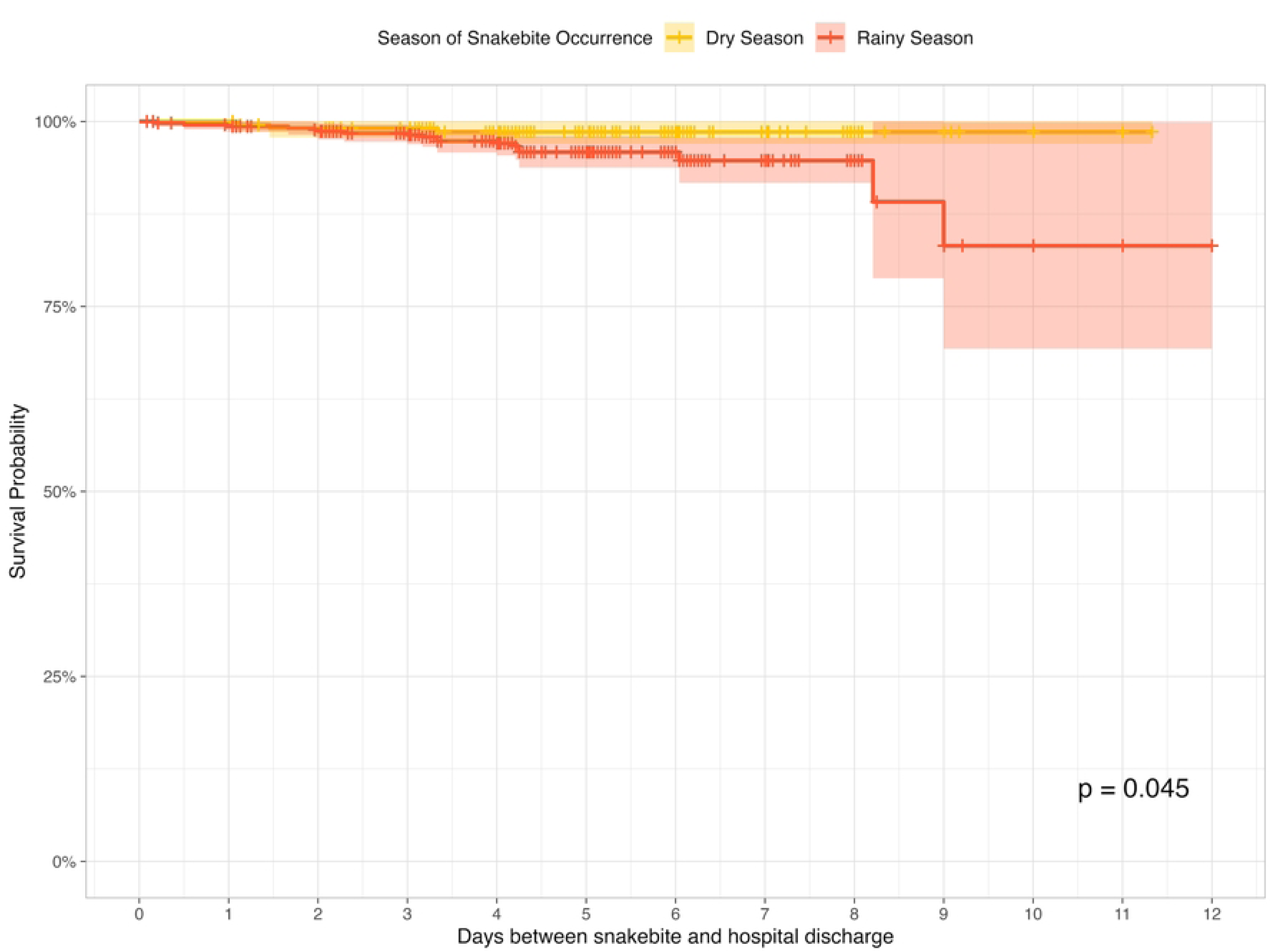
Cumulative survival probability by season of snakebite from bite to hospital discharge

Kaplan-Meier curves were also produced for the two other periods. When considering time from hospital arrival to discharge, all the same covariates remained significantly associated with cumulative survival probability: antivenom administration (p < 0.0001), antivenom cost (p < 0.0001), time from hospital admission to antivenom administration (p < 0.0001), time from bite to hospitalization (p = 0.014), and season of snakebite (p = 0.049) (Figures S9-S13). Other covariates were not significantly associated with cumulative survival probability from hospital arrival to discharge (Figures S14-S19). No covariates demonstrated a significant association with cumulative survival probability from antivenom administration to hospital discharge (Figures S20-S29); notably, this analysis only included those who received antivenom.

Subsequent Cox proportional hazards modelling revealed important predictors of survival, controlling for confounding factors. Because there was multicollinearity between the three antivenom variables (namely, antivenom administration, antivenom cost, and time to antivenom administration), we opted for three different Cox proportional hazards models, each with one of these variables. Figures 7-9 describe model results for these three models.

In the first model, the time between bite and hospitalization significantly impacted survival. Those who took four hours or more to arrive at the hospital after being bitten were more likely to die at hospital discharge compared to those who arrived in less than four hours (hazard ratio (HR) = 5.87, 95% CI = 1.15-29.95). Further, those who were not given antivenom were more likely to die at hospital discharge than those who were given antivenom (HR = 10.07, 95% CI = 3.89-26.07). Finally, being part of the 5 to 9 age group appeared protective against death (HR = 0.19, 95% CI = 0.04-0.98), compared to the 0 to 4 age group (Figure 7).

**Fig 7.**
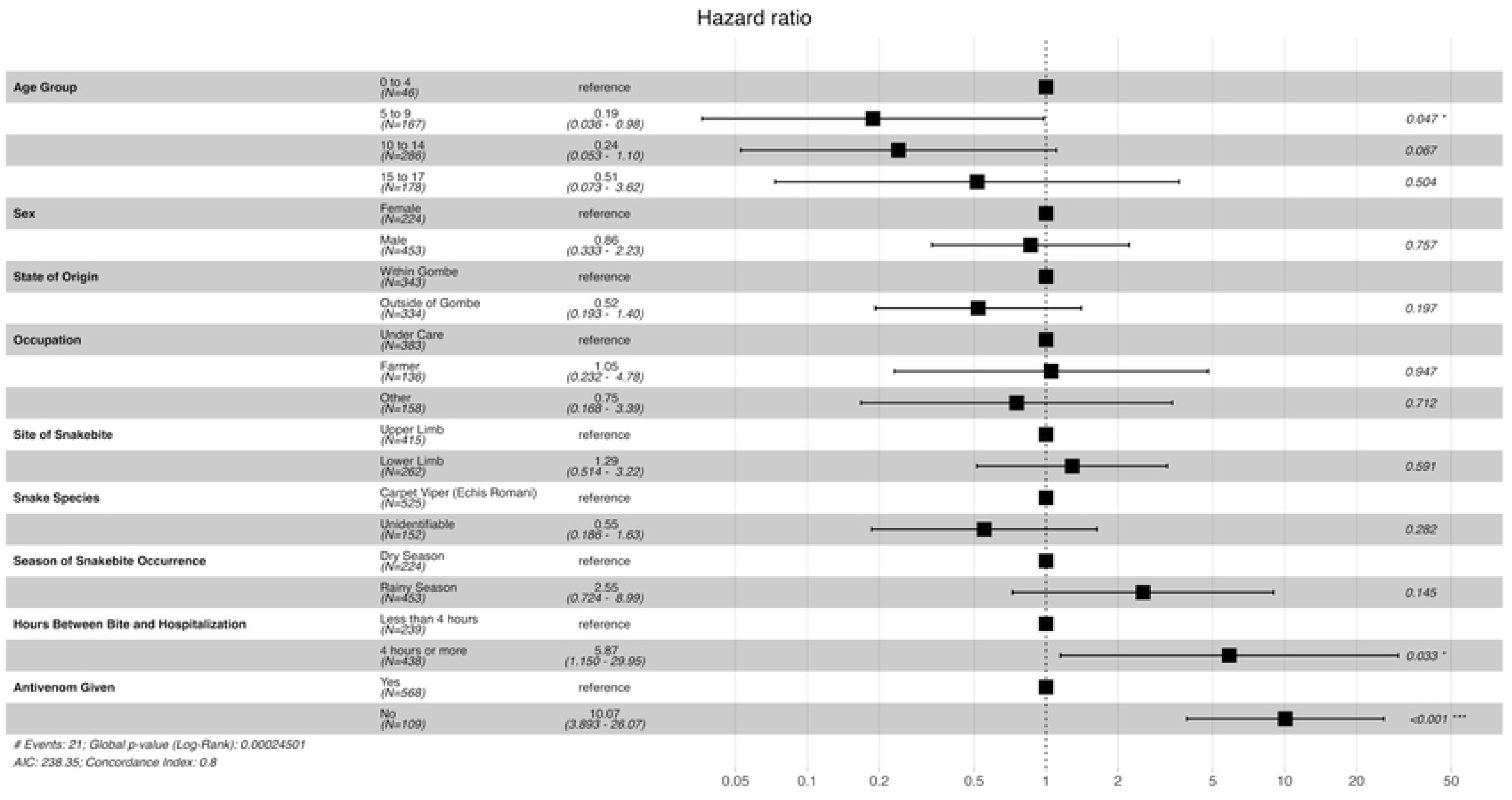
Cox proportional hazards model for survival between snakebite and hospital discharge (antivenom administration)

In the second model, the directionality of the association between variables significantly associated with the outcome remained unchanged. Like in the first model, those who took four hours or more to arrive at the hospital after being bitten were more likely to die at hospital discharge compared to those who arrived in less than four hours (HR = 5.84, 95% CI = 1.14-29.85). Additionally, those in the 5 to 9 year-old age group were less likely to die (HR = 0.19, 95% CI 0.04-0.97) compared to those in the 0 to 4 year-old age group. Finally, those who were given free antivenom or paid antivenom were less likely to die at hospital discharge than those were not given antivenom at all (HR = 0.09, 95% CI = 0.03-0.30 for free antivenom; HR = 0.11, 95% CI = 0.04-0.34 for paid antivenom) (Figure 8).

**Fig 8.**
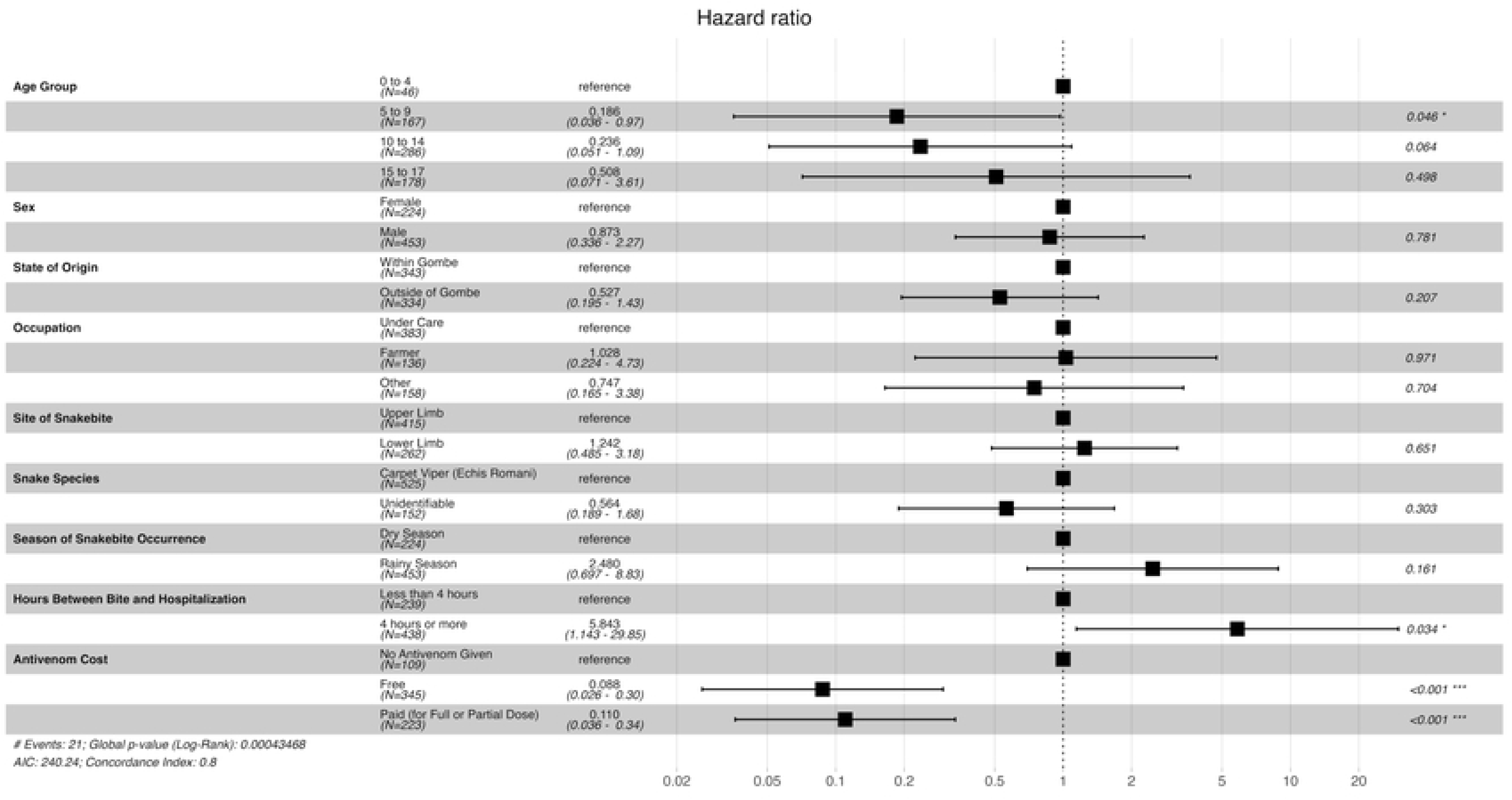
Cox proportional hazards model for survival between snakebite and hospital discharge (antivenom cost)

Finally, in the third model, like in the first and second models, those who took four hours or more to arrive at hospital after being bitten were more likely to die at hospital discharge compared to those who arrived in less than four hours (HR = 5.86, 95% CI = 1.15-29.90). Similarly, patients in the 5 to 9 age group had better survival compared to the 0 to 4 age group (HR = 0.18, 95% CI 0.04-0.96). Further, those who were given antivenom within one hour or after one hour or more were less likely to die at hospital discharge than those were not given antivenom at all (HR = 0.05, 95% CI = 0.01-0.45 for antivenom in less than one hour; HR = 0.11, 95% CI = 0.04-0.29 for paid antivenom) (Figure 9).

**Fig 9.**
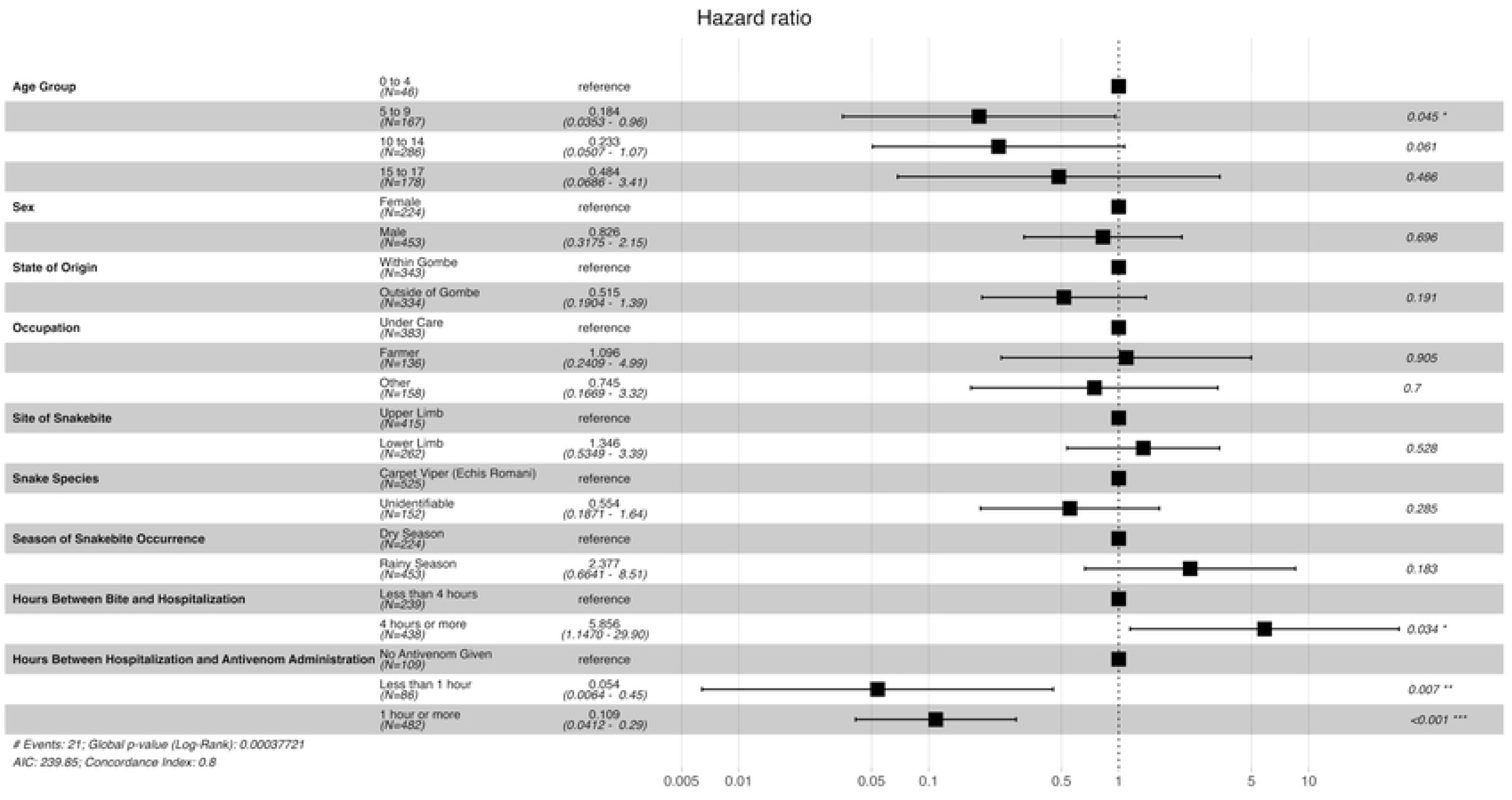
Cox proportional hazards model for survival between snakebite and hospital discharge (time to antivenom administration)

Similar Cox proportional hazards models were created using a time period from hospital arrival to discharge (Figures S30-S32). Two variables, age and occupation, were removed from these models due to a violation of the proportional hazards assumption. The significance and directionality of the relationships between time from bite to hospitalization and the three antivenom variables remained consistent in these models. Notably, Cox proportional hazards models were not generated for the time from antivenom administration to hospital discharge due to numerous violations of the proportional hazards assumptions, which prevented the models from converging.

## Discussion

This study presents survival data from 723 paediatric snakebite cases managed at SBTRH between January and December 2024. With 21 documented in-hospital deaths, the findings offer valuable insights into predictors of mortality among children and adolescents in snakebite-endemic regions of Northeastern Nigeria. This subset of paediatric patients represents a significant portion of the annual caseload at SBTRH Kaltungo, a high-burden treatment centre, thus providing a focused understanding of paediatric vulnerability and response to SBE.

Our results reveal that while most paediatric patients survived their envenomation (97%), there were key factors significantly associated with the likelihood of death. Time from bite to hospital arrival consistently emerged as a strong predictor of mortality. Patients who presented four or more hours after the bite were nearly six times more likely to die than those who arrived earlier; a finding that remained significant across all three cox proportional hazards models for the time period from snakebite to hospital discharge. This aligns with existing literature emphasizing the importance of rapid access to medical care following envenomation (11,18,19,20,21). Delays in presentation may reflect barriers such as poor transportation infrastructure, reliance on traditional healers, low awareness of the urgency of treatment, and socioeconomic constraints, particularly in rural settings. These findings underscore the need for strengthened community education, improved referral systems, and investment in emergency transport infrastructure to reduce time to care.

Antivenom administration was also strongly associated with improved survival across all analyses. Antivenom availability, timing, and cost were some of the strongest predictors of survival. Patients who did not receive antivenom were significantly more likely to die compared to those who did (Adjusted hazard ratio (aHR) = 10.07, 95% CI: 3.89–26.07, *p* < 0.001). The timing of administration also played a critical role: patients who received antivenom within one hour of hospital admission had a 95% lower hazard of death (aHR = 0.05, 95% CI: 0.01–0.45, *p* < 0.001), while those treated after one hour also experienced significantly reduced mortality (aHR = 0.11, 95% CI: 0.04–0.29, *p* < 0.001), compared to those who did not receive antivenom at all. Cost was similarly influential. Patients who received free antivenom had a 91% lower risk of death than those who received none (aHR = 0.09, 95% CI: 0.03–0.30, *p* < 0.001), and those who paid still showed substantial protection (aHR = 0.11, 95% CI: 0.04–0.34, *p* < 0.001).

Indeed, over the years, antivenom has been considered the cornerstone of envenomation treatment (2, 24, 25, 26), and these results reinforce its essential role in reducing mortality. Notably, patients who did not receive antivenom were typically affected by stockouts, limited supply, or financial barriers, underscoring persistent access challenges. Kaplan-Meier survival curves further confirmed these associations (*p* < 0.001), consistently showing higher cumulative survival among those who received antivenom, regardless of timing or cost. These findings highlight the urgent need for policies that ensure timely, universal, and affordable access to antivenom, particularly in high-burden, resource-limited settings.

Age group also demonstrated a significant relationship with survival. Patients aged 5 to 9 had a markedly reduced hazard of death compared to those aged 0 to 4 (aHR **=** 0.19, 95% CI = 0.04–0.98, p = 0.047), suggesting that very young children may be more vulnerable to the systemic effects of venom due to smaller body size or differences in physiological response (16,17, 18, 27). However, this warrants further investigation. Similarly, while sex, occupation, and site of bite were not significantly associated with survival in this cohort, the observed trends still warrant further exploration in larger or multicenter studies to confirm or refute these patterns. In addition, some covariates, such as age and occupation, had to be excluded from the cox proportional hazards models evaluating survival from admission to discharge due to violation of the proportional hazards assumptions. This limitation may have constrained the interpretation of certain subgroup effects in that time frame. Targeted interventions and heightened clinical vigilance may therefore be necessary for very young children in areas endemic to snakebites.

Seasonality was another key factor associated with survival. In descriptive analyses, patients bitten during the rainy season had worse outcomes compared to those bitten in the dry season (p = 0.045), with an average case fatality rate of 3.5% during the rainy months compared to 1.8% in the dry season a finding that may reflect increased snake activity, heightened exposure due to farming, or environmental conditions that delay access to care. Similar seasonal patterns have been observed in other regional and global snakebite studies (28, 29, 30). These findings suggest the need for targeted seasonal preparedness and resource planning, such as stocking antivenom and conducting community education during the high-risk months.

Interestingly, snake species identification was not significantly associated with survival despite the fact that most patients (77%) were bitten by carpet vipers (Echis romani), a species known for its potent hemotoxic venom (12, 31,32). Despite this, statistical analysis revealed no significant difference in survival based on snake species (p=0.831), suggesting that species identification alone may not be a reliable predictor of outcome in this context. As in previous studies (11, 23), a substantial number of patients (22%) were unable to identify the snake species. This highlights the need for improved community education on the importance of snake identification, which can aid in guiding appropriate antivenom use and potentially improve treatment outcomes.

Notably, no covariates were found to be significantly associated with the cumulative survival probability from the time of antivenom administration to hospital discharge. This may suggest that, once antivenom is administered, a patient’s likelihood of survival is largely established. Alternatively, the absence of significant associations may reflect the limited statistical power within this subgroup of patients who received antivenom. Regardless, the finding underscores that the most critical window for reducing mortality lies in the period preceding antivenom administration, highlighting the importance of early intervention and rapid access to treatment.

In addition to descriptive and survival analyses, this study leveraged Cox proportional hazards modeling to identify adjusted predictors of mortality. Although all models consistently showed the protective effect of early hospital presentation and antivenom access, multicollinearity among antivenom-related variables limited the interpretability when all three models were included simultaneously. This challenge highlights the statistical complexity of modeling closely related treatment variables, especially when they reflect both clinical decision-making and patient severity. Nevertheless, this modeling approach highlights the utility of survival analysis in clinical snakebite research and offers a potential foundation for future risk stratification tools in future studies.

Our study responds to the growing recognition of the need for targeted paediatric snakebite research by addressing key methodological limitations in the existing literature. Most studies to date rely on simple descriptive or cross-sectional analyses, which fail to account for when deaths occur or how risk evolves over time. In contrast, our research pioneers the application of survival analysis techniques such as the Kaplan-Meier estimator and cox proportional hazards regression, a facility that has consistently managed snakebite cases from rural northeastern Nigeria, parts of northwestern and north central Nigeria, as well as neighboring regions of Cameroon and the Republic of Chad, with a notably high recovery rate (11). These approaches enable a more detailed and time-sensitive understanding of mortality risk (22), ultimately supporting the development of evidence-based strategies for prevention, triage, and clinical management tailored to vulnerable pediatric populations.

While our study demonstrates the potential for advanced statistical modeling in predicting snakebite outcomes, it also reveals critical limitations. Prominent among these is the single-centre nature of the data, which may limit generalizability to other regions with differing health infrastructure, snake species prevalence, and cultural practices. Additionally, despite using a relatively large paediatric sample for snakebite research, the number of deaths (n = 21) was small, limiting statistical power. Furthermore, certain potentially influential variables such as envenomation severity scores, presence of systemic bleeding or neurological symptoms, and initial vital signs were not available in the dataset. These factors could further refine predictive models and clinical algorithms.

Future studies should focus on multicentre collaborations that standardize data collection and reporting. Inclusion of clinical markers of severity and real-time physiological data would enhance the development of more robust prognostic tools. Moreover, integrating these tools into clinical workflows, such as electronic decision-support systems, has the potential to improve outcomes through timely triage and intervention.

## Conclusions

This study identifies delayed hospital presentation, lack of antivenom administration, and, potentially, very young age as key predictors of in-hospital mortality among paediatric snakebite patients in Northeastern Nigeria. These findings underscore the need for urgent public health interventions, including the strengthening of antivenom supply chains, eliminating financial barriers to treatment, and the implementation of targeted educational campaigns aimed at early care-seeking behaviour.

Our study also demonstrates the feasibility and value of using survival analysis techniques to understand predictors of death in paediatric envenomation and lays the foundation for future development of clinical decision-support models. We recommend disseminating these results through regional health networks and academic forums, and urge policymakers to prioritize the provision of free and timely antivenom as a central strategy to reduce snakebite mortality in vulnerable populations.

Future studies should focus on prospective, multicentre studies that incorporate standardized data collection and clinical severity markers to refine and validate predictive models and inform evidence-based clinical management protocols.

## Data Availability

The data that support the findings of this study are not openly available due to reasons of sensitivity and are available from the corresponding authors upon reasonable request. Data are located in controlled access data storage at the Snakebite Treatment and Research Hospital, Kaltungo.

## Acknowledgements

We would like to acknowledge the key guidance of the Gombe State Hospital Management Board and the Gombe State Ministry of Health.

## Supporting information captions

**Figure S1.** Overall survival probability from hospital arrival to discharge

**Figure S2.** Overall survival probability from antivenom administration to hospital discharge

**Figure S3.** Cumulative survival probability by age group from bite to hospital discharge

**Figure S4.** Cumulative survival probability by age group from bite to hospital discharge

**Figure S5.** Cumulative survival probability by sex from bite to hospital discharge

**Figure S6.** Cumulative survival probability by site of snakebite from bite to hospital discharge

**Figure S7.** Cumulative survival probability by snake species from bite to hospital discharge

**Figure S8.** Cumulative survival probability by state of origin from bite to hospital discharge

**Figure S9.** Cumulative survival probability by antivenom administration from hospital arrival to discharge

**Figure S10.** Cumulative survival probability by antivenom cost from hospital arrival to discharge

**Figure S11.** Cumulative survival probability by hours between hospital admission and antivenom administration from hospital arrival to discharge

**Figure S12.** Cumulative survival probability by hours between bite and hospitalization from hospital arrival to discharge

**Figure S13.** Cumulative survival probability by season of snakebite from hospital arrival to discharge

**Figure S14.** Cumulative survival probability by age group from hospital arrival to discharge

**Figure S15.** Cumulative survival probability by occupation from hospital arrival to discharge

**Figure S16.** Cumulative survival probability by sex from hospital arrival to discharge

**Figure S17.** Cumulative survival probability by site of snakebite from hospital arrival to discharge

**Figure S18.** Cumulative survival probability by snake species from hospital arrival to discharge

**Figure S19.** Cumulative survival probability by state of origin from hospital arrival to discharge

**Figure S20.** Cumulative survival probability by age group from antivenom administration to hospital discharge

**Figure S21.** Cumulative survival probability by antivenom cost from antivenom administration to hospital discharge

**Figure S22.** Cumulative survival probability by hours between hospital admission and antivenom administration from antivenom administration to hospital discharge

**Figure S23.** Cumulative survival probability by hours between bite and hospitalization from antivenom administration to hospital discharge

**Figure S24.** Cumulative survival probability by occupation from antivenom administration to hospital discharge

**Figure S25.** Cumulative survival probability by season of snakebite from antivenom administration to hospital discharge

**Figure S26.** Cumulative survival probability by sex from antivenom administration to hospital discharge

**Figure S27.** Cumulative survival probability by site of snakebite from antivenom administration to hospital discharge

**Figure S28.** Cumulative survival probability by snake species from antivenom administration to hospital discharge

**Figure S29.** Cumulative survival probability by state of origin from antivenom administration to hospital discharge

**Figure S30.** Cox-proportional hazards model for survival between hospital arrival and discharge (antivenom administration)

**Figure S31.** Cox-proportional hazards model for survival between hospital arrival and discharge (antivenom cost)

**Figure S32.** Cox-proportional hazards model for survival between hospital arrival and discharge (time to antivenom administration)

## Notes

### Competing Interest Statement

The authors have declared no competing interest.

### Funding Statement

Aashna Uppal acknowledges the receipt of studentship awards from the Health Data Research UK-The Alan Turing Institute Wellcome PhD Programme in Health Data Science (Grant Ref: 218529/Z/19/Z). No other author received funding for this work.

### Author Declarations

Ethical clearance was obtained from the Gombe State Health Research Committee, part of the Ministry of Health (Ref: MOH/ADM/621/V.1/497), and necessary approvals were obtained from the Gombe State Hospital Services Management Board. Through this approval process, the requirement for individual patient consent was waived.

